# Molecular Neuropathology and Cerebrospinal Fluid Diagnostic Biomarkers of SARS-Cov2 Infection in Central Nervous System– A Scoping Review Protocol

**DOI:** 10.1101/2023.03.17.23287405

**Authors:** Victor Meza Kyaruzi, Emmanuel Mduma, Arsene Daniel Nyalundja, Soham Bandyopadhyay, Larrey Kasereka Kamabu, Bydaa Atron, Mugenyi Nathan, Jeremiah Oluwatomi Itodo Daniel, Zobidah Yousif Elamin, Boniphace Barnabas Marwa, Rajab Msemo, Ahmed Naeem, Tumusifu Manegabe jean de Dieu, Tarun Suvari Kumar, Ngepgou Beckline Tazoah, Ugwoke Franklin Chiazo, Samuel Oreoluwa David, Yves Jacket Nsavyimana, Constansia Anselim Bureta, Nicephorus Rutabasibwa, Laurent Lemeri Mchome, Emnet Tesfaye Shimber, Abenezer Tirsit, Sayoki Mfinanga, Getaw Worku Hassen, Osama Abdellaziz, Amos Mwakigonja

## Abstract

**Introduction:** Despite the broad spectrum of neurological symptomatic manifestation in COVID19 patients, the brain tissue susceptibility and permissiveness to SARS-Cov2 infection is yet uncertain. This critical appraisal aims at bridging the gap by consolidating the body of evidence for meticulous evaluation of molecular neuropathological pathways and CSF diagnostic signatures of SARS-Cov2 infection in the central nervous system (CNS) that will underpin further strategic approach for neuroprotection and treatment of neurological COVID19

**Methods and Analysis:** We have developed the protocol of this review according to the provisions of Joanna Briggs Institute Reviewer’s Manual for Evidence Synthesis, 2015 and Arksey and O Malley’s Methodological Framewotk, 2005. The articles for this review will be sourced from several electronic databases including EMBASE, PubMed, Scopus, Web of Science (WOS), Cochrane, Crossref Metadata and Semantic scholar. Herein we generated the search strategy using the medical subject headings [ MeSH Terms], term in all field bibliography at all permutations in conjunctions with boolean operators

**Ethical Clearance and Dissemination plan:** Herein the review will not involve the human participants henceforth the ethical clearance approval is not applicable. We will disseminate the final findings of this review to scientific conferences at local and international level. The manuscript for final findings will be published on reputable journal of neuroscience.

## INTRODUCTION

The classical pathogenic pathway of SARS-Cov2 infection has been elucidated in the respiratory system where viral host tropism is prominently elaborated by expression and upregulation of angiotensin converting enzyme 2 (ACE2) receptors on the apical membrane of respiratory epithelium (1,2). However the wide distribution of ACE2 receptors across multiple organs including the heart, kidneys, gastrointestinal, pancreas and severe complications such as ischaemic stroke, haemorhagic encephalopathy and posterior reversible encephalopathy syndrome with epileptic seizures have been reported in more than one-third of the COVID19 hospitalized patients (3,4).

Despite the broad spectrum of neurological symptomatic manifestation, the neuropathological pathway is yet uncertain, the brain tissue susceptibility and permissiveness to SARS-Cov2 has not been elucidated. Whether the pathophysiology and constellation of symptoms is concealed within the direct viral neuroinvasion or systemic viremia and inflammatory response syndrome is still indeterminate (5).

The idiosyncratic mechanisms of how the virus and cytokines invade and penetrate the brain blood barrier (BBB) surrounding the brain tissue is undefined. However both preclinical and human studies have described some hypothetical neuroinvasion pathways such as retrograde axonal transportation from peripheral nervous system to the central nervous system, transcribrial route, ocular surface and haematogenous route. Several cell surface receptors such as ACE2, transmembrane protease serine 2 (TMPRSS2) and P2X7 have been identified as the culprit of SARS-Cov2 neuropathogenesis (6,7).

Severity of neuronal damage and neuroinflammation may be detected by the level of biomarker concentration in both serum and CSF fluid, however the sensitivity and specificity of these biomarkers is still debatable. Elevated levels of neurofilament light chain protein (NfL) and glial fibrillary acidic protein (GFAp) delineate the ongoing acute neuronal injury and astrocytic damage respectively in adjunct with detection of other biomarkers such as T tau and viral RNA. Despite of their diagnostic assays significant concentration variation for encephalitis, encephalopathy and acute disseminated encephalomyelitis (ADEM) has been elaborated (8–11).

To our knowledge this critical appraisal will bridge the gap by consolidating the body of evidence for meticulous evaluation of molecular neuropathological pathways and CSF diagnostic signatures of SARS-Cov2 infection in the central nervous system (CNS) that will underpin further strategic approach for neuroprotection and treatment of neurological COVID19.

### Review objectives

#### Primary objective

To determine the molecular neuropathological pathways of SARS-Cov2 infection in CNS

#### Secondary objectives

1. To describe the neuropathological manifestation of SARS-Cov2 infection in CNS
2. To determine the types and accuracy of CSF diagnostic biomarkers of SARS-Cov2 infection in CNS

## METHODS AND ANALYSIS

We have developed the protocol of this review according to the provisions of Joanna Briggs Institute Reviewer’s Manual for Evidence Synthesis, 2015 and Arksey and O Malley’s Methodological Framewotk, 2005 (12,13). A scope review is considered suitable for the fitness of our study design because of outstanding paucity for strong evidence to unfold the uncertainty about the context and concepts of molecular neuropthophysiology and CSF diagnostic signatures of SARS-Cov2 infection in CNS.

Herein the methodology approach for this protocol is adopted according to the five stages of Arksey and O’Malley’s Framework

### Stage 1. Identifying the research question

The research question in our protocol has been described in alignment with the Participants, Concept and Context (PCC) strategy.

#### Participants

All human subjects at any age diagnosed with COVID19 and presenting with neurological manifestation will be evaluated in this review

#### Concept

We will evaluate the molecular neuropathological pathways and CSF diagnostic biomarkers of SARS-Cov2 infection in CNS.

#### Context

In this review we will critically appraise all existing primary sources of evidence in the broad context of neuropathogenesis of COVID19 evaluating more on the theories of molecular susceptibility and permissiveness of SARS-Cov2 tropism in the brain and spinal cord parenchyma. The type of surface receptors expressed and upregulated for viral neuroinvasion, molecular pathways involved in axonal injury and astrocytic damage, the cellular metabolic and inflammatory response following the neuroinvasion of the virus in CNS tissue. The types and diagnostic accuracy of CSF biomarkers of SARS-Cov2 infection in CNS.

### Stage 2: Identifying relevant studies

#### Information source and search strategy

##### Information source

The articles for this review will be sourced from several electronic databases including EMBASE, PubMed, Scopus, Web of Science (WOS), Cochrane, Crossref Metadata and Semantic scholar.

##### Search strategy

Herein we generated the search strategy using the medical subject headings [ MeSH Terms], term in all field bibliography at all permutations in conjunctions with boolean operators such as AND or OR.

Neuropathology [MeSH terms] OR Molecular Neuropathology [MeSH Terms] AND CSF diagnostic biomarkers [MeSH Terms] AND COVID19 OR SARS-Cov2. The results from PubMed electronic database are presented in **table 1** on appendix section.

##### Eligibility criteria

The eligibility criteria was constructed based on the PCC strategy as the operation blueprint to guide the reviewers for appropriate article selection

###### Participants

Human subjects at any age

###### Concept

All articles reporting on molecular neuropathological pathways or CSF diagnostic biomarkers of SARS-Cov2 infection in CNS will be included.

###### Context

Articles reporting on molecular neuropathogenesis theories entailing molecular susceptibility, permissiveness and invasion of the SARS-Cov2/COVID19 in the CNS parenchyma. The type of surface receptors such as ACE2, TMPRSS2, P2X7which are implicated in the pathogenesis of CNS SARS-Cov2 infections and symptom manifestations. The types of molecular pathways implicated for axonal injury and astrocytic cellular damage, associated metabolic derangements and systemic inflammatory response as the culprits of viral neuroinvasion to CNS tissue.

All articles reporting on types, sensitivity and specificity of CSF diagnostic biomarkers of SARS-Cov2 infection in CNS such as neurofilament light chain protein (NfL), glial fibrillary acidic protein (GFAp), T tau, P tau and Viral RNA.

### Stage 3: Study selection

Two reviewers VMK and EM will upload the sourced articles to Covidence software and independently screen the articles based on eligibility criteria. The screening process will be blinded and achieved in two stages, phase 1 will comprise of screening the abstracts and phase 2 will be achieved by screening of full text any conflict encountered upon screening will be resolved by consensus between the parties or adjudication by a third reviewer ADN. All results will presented using the Preferred Reporting Items for Systematic Reviews and Meta-analysis for flow diagram for screening process shown in **figure 1**.

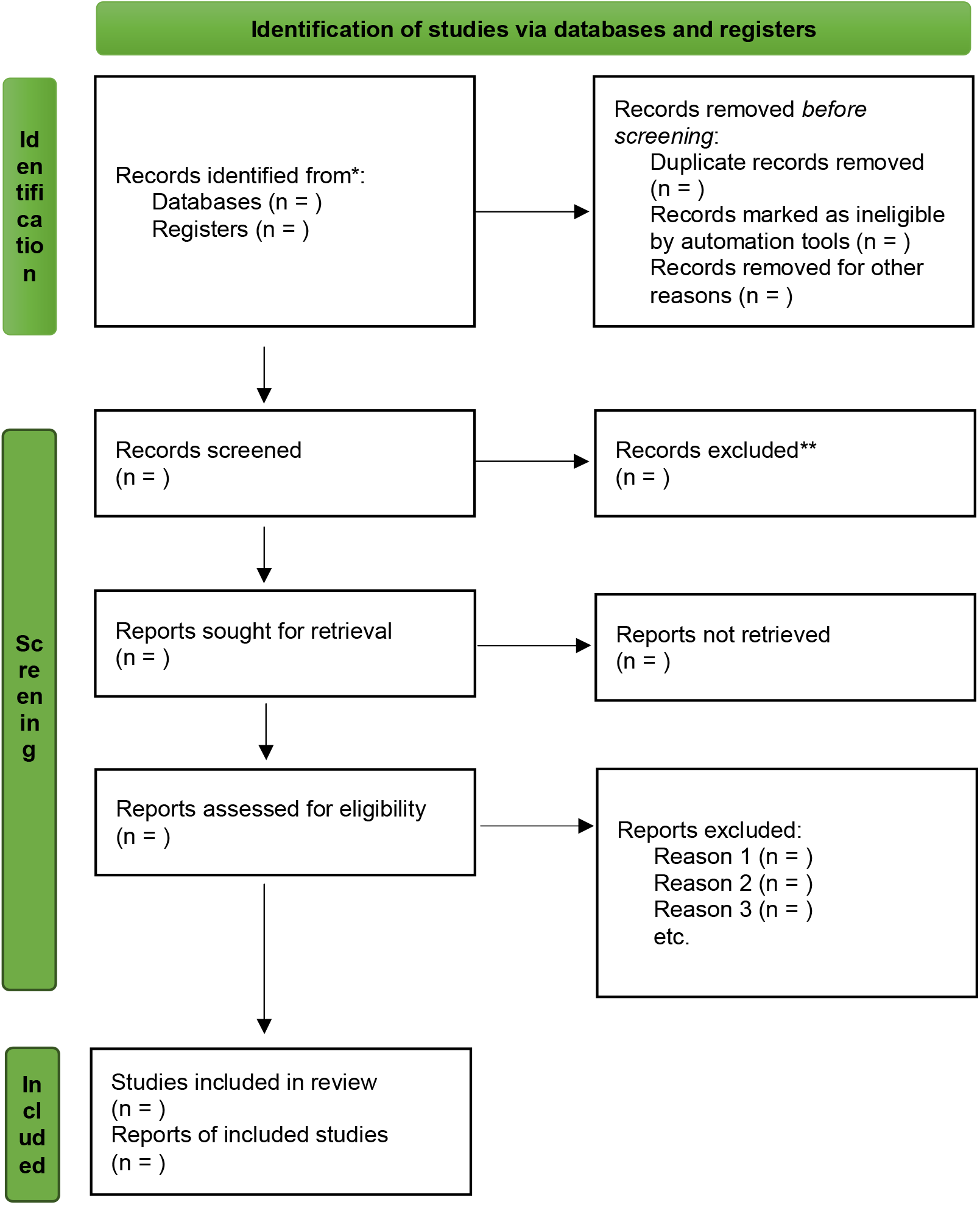
Preferred Reporting Items for Systematic Reviews and Meta-analysis for flow diagram for screening process shown in figure 1.

### Stage 4: Charting the data

The data items from selected articles will be extracted using the Cochrane Effective Practice and Organization of Care (EPOC) data collection form for systematic review (14). The reliability and validity of this tool will preliminarily tested on 10 randomly selected articles, the consistency acceptability will be determined using the Cronbach’s alpha and Kudar Richardson, the threshold value for acceptability will be optimized at ≥ 0.7(15). The data item to be extracted are enumerated in the **table 2** below

**Table 2:** Items to be extracted.

| **Table 2: Items to be extracted** |
| --- |
| **Original article information**<br>a) First Author<br>b) Year of publication<br>c) Country of origin<br>d) Aims<br>e) Sample size (n)<br>Study population and sample size<br>Methods<br>**Outcomes purporting the PCC question**<br>a) Participants : Human subjects , sex , age , patients infected with COVID19<br>b) Concept: Types of molecular neuropathological pathways , types of CSF diagnostic biomarkers<br>c) Context : Types of receptors such as ACE2 , TMPRSS2 , P2X7 and other cellular transduction pathways .The specific CSF diagnostic biomarkers for axonal injury , astrocytic cellular damage or viral neuroinvasion such neurofilament light chain protein (NfL) , glial fibrillary acidic protein (GFAP) , T tau , P tau and Viral RNA. |

### Stage 5: Collating, summarizing and reporting of results

All extracted data will be presented on matrix tables, variables such as first author, year of publication, sample size, measurements of central tendency and dispersion, molecular pathway, receptors and CSF biomarkers will be outlined. The result findings will be summarized in sections in order to generate the specific and comprehensive deductions.

### Risk of Bias (ROB) assessment

The ROB will be evaluate using the ROB tool Version 2 for RCT studies (16) and Risk of Bias in Non-randomized Studies of Interventions (ROBINS-I) for NRSI designs and the overall ROB will be rated as low risk, moderate risk, serious risk and critical risk(17).

### Strength of body of evidence

We will evaluate the strength and external applicability of the evidence findings of this review using the Grading of Recommendation, Assessment, Development and Evaluation (GRADE) protocol. The quality levels of evidence will be classified as high, moderate, low and very low level of evidence (LOE) according to the GRADE provisions(18,19).

### Ethical consideration and Dissemination

Herein the review will not involve the human participants henceforth the ethical clearance approval is not applicable. We will disseminate the final findings of this review to scientific conferences at local and international level.The manuscript for final findings will be published to reputable journal of neuroscience.

## Data Availability

All data produced in the present work are contained in the manuscript

## DECLARATION

### Contribution of Authors

Victor Meza Kyaruzi and Emmanuel Mduma prepared the full text of this protocol and all other authors reviewed and contributed their expert opinions.

Concept and Context: All authors

Drafting of Manuscript: Victor Meza Kyaruzi, Emmanuel Mduma and Daniel Nyalundja Literature Search: Victor Meza Kyaruzi and Emmanuel Mduma

Critical revision of the manuscript for intellectual content: All authors

Supervision: Prof Abenezer Tirsit, Prof Sayoki Mfinanga, Prof Getaw Worku Hassen, Prof Osama S Abdelaziz and Prof Amos Mwakigonja.

## Appendix

**Table 1:** PubMed Search Strategy. <u>Advanced Search Results - PubMed</u> (nih.gov)

| Search | Query |
| --- | --- |
| #6 | Search: **#1 OR #2 AND #3 AND #4 OR #5**<br>((("neuropathology"[MeSH Terms] OR (("molecular"[All Fields] OR "moleculars"[All Fields]) AND "neuropathology"[MeSH Terms]))) AND ((("CSF"[All Fields] AND ("diagnosis"[MeSH Terms] OR "diagnosis"[All Fields] OR "diagnostic"[All Fields] OR "diagnostical"[All Fields] OR "diagnostically"[All Fields] OR "diagnostics"[All Fields]))) AND "biomarkers"[MeSH Terms]) AND "covid 19"[MeSH Terms]) OR "SARS-Cov2"[All Fields]<br>**Translations**<br>**Neuropathology[MeSH Terms]:** "neuropathology"[MeSH Terms]<br>**Molecular:** "molecular"[All Fields] OR "moleculars"[All Fields]<br>**Neuropathology[MeSH Terms]:** "neuropathology"[MeSH Terms]<br>**Diagnostic:** "diagnosis"[MeSH Terms] OR "diagnosis"[All Fields] OR "diagnostic"[All Fields] OR "diagnostical"[All Fields] OR "diagnostically"[All Fields] OR "diagnostics"[All Fields]<br>**biomarkers[MeSH Terms]:** "biomarkers"[MeSH Terms]<br>**COVID19[MeSH Terms]:** "covid-19"[MeSH Terms] |
| #5 | Search: **SARS-Cov2**<br>"SARS-Cov2"[All Fields] |
| #4 | Search: **COVID19[MeSH Terms]**<br>"covid 19"[MeSH Terms]<br>**Translations**<br>**COVID19[MeSH Terms]:** "covid-19"[MeSH Terms] |
| #3 | Search: **CSF Diagnostic biomarkers[MeSH Terms]**<br>("CSF"[All Fields] AND ("diagnosis"[MeSH Terms] OR "diagnosis"[All Fields] OR "diagnostic"[All Fields] OR "diagnostical"[All Fields] OR "diagnostically"[All Fields] |
|  | OR "diagnostics"[All Fields])) AND "biomarkers"[MeSH Terms]<br>**Translations**<br>**Diagnostic:** "diagnosis"[MeSH Terms] OR "diagnosis"[All Fields] OR "diagnostic"[All Fields] OR "diagnostical"[All Fields] OR "diagnostically"[All Fields] OR "diagnostics"[All Fields]<br>**biomarkers[MeSH Terms]:** "biomarkers"[MeSH Terms] |
| #2 | Search: **Molecular Neuropathology[MeSH Terms]**<br>("molecular"[All Fields] OR "moleculars"[All Fields]) AND "neuropathology"[MeSH Terms]<br>**Translations**<br>**Molecular:** "molecular"[All Fields] OR "moleculars"[All Fields]<br>**Neuropathology[MeSH Terms]:** "neuropathology"[MeSH Terms] |
| #1 | Search: **Neuropathology[MeSH Terms]**<br>"neuropathology"[MeSH Terms]<br>**Translations**<br>**Neuropathology[MeSH Terms]:** "neuropathology"[MeSH Terms] |

